# The Para-ISS Dural Space Enhancement Sign: A Novel MRI Finding Independently Associated With Intracranial Atherosclerotic Burden

**DOI:** 10.64898/2026.11.04.26367240

**Authors:** Shaoliang Zeng, Daiying Lin, Kai Tang, Fuxin Zeng, Xufeng Zheng, Mu Li, Jinyu Lin, Baoquan Huang, Sanmei Zhan, Shaoxian Chen, Zhuozhi Dai

## Abstract

**Background:** No dural enhancement sign centered on the inferior sagittal sinus (ISS) has been characterized. We observed a frequent, previously undescribed finding—para-ISS dural space enhancement (PISE)—at the free inferior margin of the falx cerebri.

**Objective:** To define PISE, determine its detection rate in a large cross-sectional cohort, and characterize its correlates, in particular whether the pattern of association is compatible with an atherosclerosis-related dural mechanism.

**Methods:** Consecutive contrast-enhanced intracranial vessel-wall MRI (VWI) examinations (May 2015–December 2025) were reviewed; 835 of 875 examinations were evaluable, corresponding to 811 patients. The first outcome-evaluable examination for each patient was used for all primary analyses. PISE was defined as dural enhancement strictly at the falx free margin adjacent to the ISS, exceeding the ISS vascular lumen. Forty-seven demographic, clinical-history, and imaging variables were tested univariately with Benjamini–Hochberg false discovery rate (FDR) correction; multivariable logistic regression with patient-grouped 5-fold cross-validation was triangulated with LASSO and random forest. Dose–response across plaque grades was assessed with the Cochran–Armitage trend test, and circulation-specific associations were compared in models adjusted for age and sex (inter-reader Cohen’s κ=0.93).

**Results:** PISE was detected in 609/811 patients (75.1%); detection rose from 50.0% (<40 years) to 81.7% (>70 years; trend p=5.6×10⁻⁶). The strongest univariate correlate was vulnerable intracranial plaque (OR=8.29, 95%CI 5.54–12.40). Detection increased across plaque grades 0/1/2 (38.7%→77.2%→90.3%; Z=11.09). Anterior-circulation plaque showed a stronger association than posterior-circulation plaque (OR=4.42 vs 2.76). Vulnerable plaque (OR=2.37), plaque grade (OR=2.84 per grade), age (OR=1.38 per SD), and inpatient status (OR=3.18) remained independently associated (patient-grouped cross-validated AUC=0.755). Moyamoya disease was inversely associated (OR=0.25). Findings were consistent across imputation and all-examination sensitivity analyses.

**Conclusions:** PISE is a frequently detected novel dural enhancement sign whose independent, graded, anatomically concordant association with intracranial atherosclerotic burden supports an atherosclerosis-related dural mechanism and warrants prospective validation.

## 1. Introduction

On contrast-enhanced intracranial vessel-wall MRI (VWI) we repeatedly observed a distinctive pattern of dural enhancement that does not correspond to any entry in published catalogues of meningeal enhancement patterns [1,2]: tubular or band-like enhancement situated precisely at the free inferior margin of the falx cerebri, centered on the inferior sagittal sinus (ISS), within the peri-ISS dural space of the falx. We termed this finding the para-ISS dural space enhancement (PISE) sign. Physiological dural enhancement, as characterized in prospective and early contrast MRI studies, is thin, linear, discontinuous, and located along the body of the falx, the tentorium, and the convexity dura [3–5]; the enhancement we describe occupies a territory that does not overlap these descriptions. We therefore sought to define PISE operationally and to determine what it signifies. Representative MR images and corresponding schematic illustrations of its anatomical location are shown in Figure 1.

**Figure 1.**
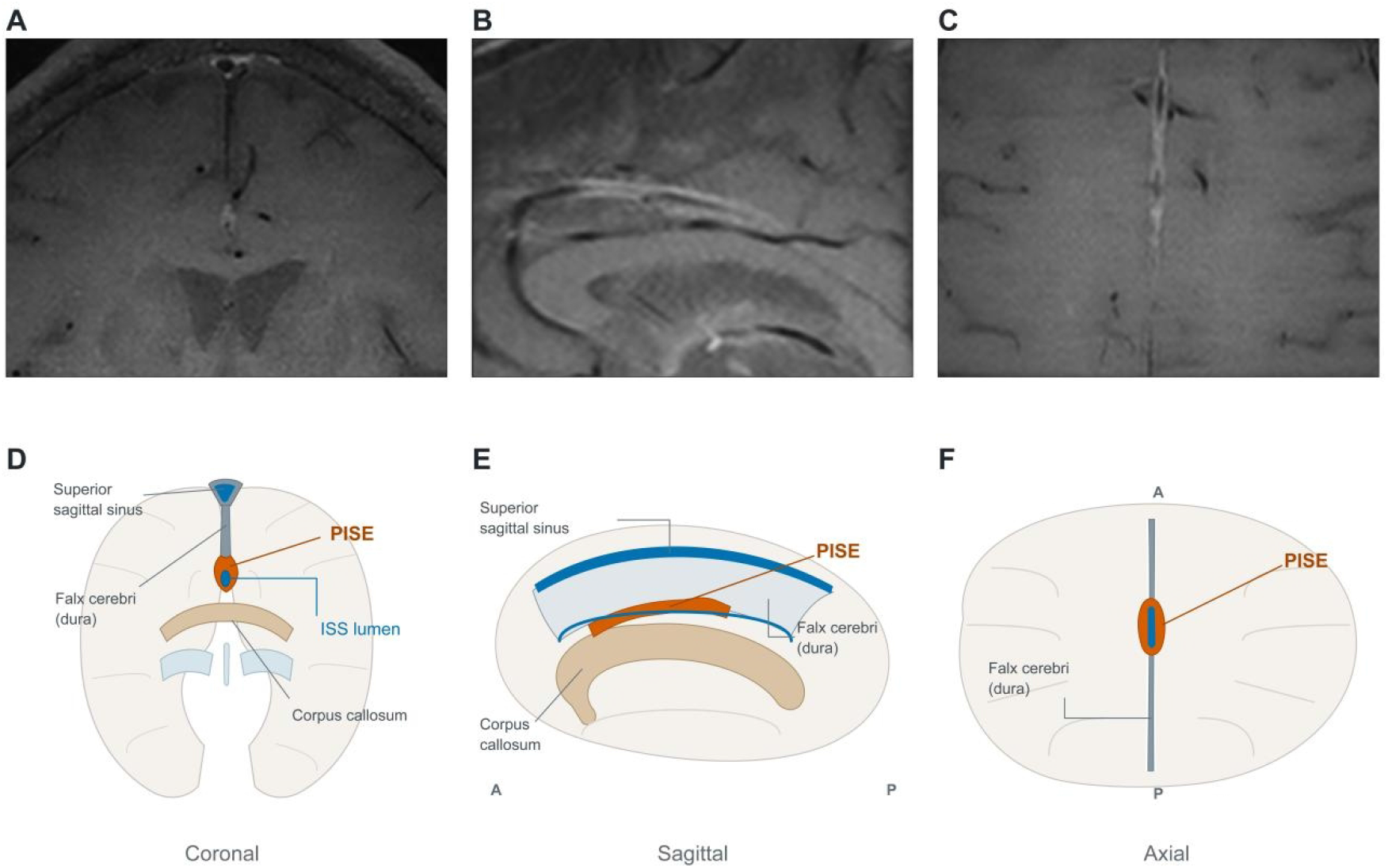
The PISE sign on post-contrast intracranial vessel-wall MRI and its schematic anatomical depiction in three orthogonal planes. (A–C) Representative PISE-positive examination in coronal (A), sagittal (B), and axial (C) planes: tubular or band-like enhancement is situated strictly at the free inferior margin of the falx cerebri, centered on the inferior sagittal sinus (ISS), exceeding the luminal contour of the sinus within the peri-ISS dural space; the finding is appreciable in all three planes. (D–F) Corresponding schematic diagrams in coronal (D), sagittal (E), and axial (F) planes illustrate the anatomical location of PISE (orange) and its spatial relationship to the ISS lumen and falx cerebri (dura); the superior sagittal sinus and corpus callosum are labeled where shown. All MR images are post-contrast black-blood vessel-wall MRI. Abbreviations: ISS, inferior sagittal sinus; MRI, magnetic resonance imaging; PISE, para-ISS dural space enhancement.

The ISS courses within the free edge of the falx cerebri, receives falcine veins and anterior pericallosal tributaries, and drains posteriorly into the vein of Galen and the straight sinus [6,7]. Thin-section contrast MRI depicts the ISS in approximately 97% of adults, with substantial morphological variability [7]; a persistent falcine sinus is a recognized variant that must be distinguished from peri-sinus dural findings [8]. Functionally, the ISS sits at a junction between the deep venous drainage of the anterior circulation and the dura that invests it, making the peri-ISS dura a candidate interface between vascular, interstitial, and meningeal compartments.

Over the past decade the dura has been re-framed from a passive covering to an active drainage and signaling interface. Meningeal lymphatic vessels were characterized in rodent dura [9,10] and visualized non-invasively in human and primate meninges by MRI [11]. Intrathecal tracer studies demonstrated cerebrospinal fluid (CSF) efflux to the parasagittal dura in humans [12], and dynamic contrast-enhanced (DCE) MRI showed that the parasagittal dural space enhances slowly with delayed washout, a kinetic distinct from venous structures [13]; cross-sectional MRI studies further showed that the parasagittal and peri-sinus dural space enlarges and its washout decreases at older age [13–15]. Histologically, lymphatic vessels accompany the dorsal dural sinuses—including the ISS—in human specimens, with larger-caliber vessels along the dorsal sinuses [16], and meningeal lymphatic circuits are conserved between mice and humans [17], although their distribution is widespread and not restricted to sinus walls [18]. Notably, a published 7-Tesla MRI atlas of the meningeal lymphatic network assessed fifteen peri-vascular and peri-sinus sites but did not include the ISS region [19]; the peri-ISS dura remains a blank spot in published imaging atlases. Several recent studies and reviews now place the parasagittal dura at the center of CSF transport, neuroimmune surveillance, and brain clearance frameworks [20–23].

Direct imaging contact points between dural enhancement and cerebrovascular disease are scarce. Delayed vessel-wall MRI in 11 patients with recent cerebral infarction showed ipsilateral delayed dural/meningeal enhancement in all five large infarctions and in none of six small infarctions or six moyamoya patients [24]; DCE-MRI studies have demonstrated impaired meningeal lymphatic function in cerebral small vessel disease [25,26], complemented by indirect diffusion-based glymphatic markers [27]; and dural thickening on FLAIR was found in 34% of a neurodegenerative cohort, largely unrelated to vascular risk factors [28]. A recent review concluded that human MRI evidence on post-stroke meningeal lymphatics remains exploratory [29]. No study has systematically characterized a dural enhancement sign on contrast-enhanced MRI in relation to intracranial atherosclerotic burden in a large cohort.

We therefore asked a descriptive-mechanistic question: in a large cross-sectional cohort of contrast-enhanced VWI examinations, what is the detection rate of PISE, what are its correlates, and does the pattern of correlates point to an atherosclerosis-related dural mechanism? Specifically, we tested whether PISE is independently associated with atherosclerotic burden (strength), whether detection rises monotonically with plaque grade (dose–response), whether the association is stronger for the anterior than the posterior circulation (anatomical correspondence with ISS drainage), and whether a non-atherosclerotic arteriopathy—moyamoya disease—behaves differently (natural control).

## 2. Methods

### 2.1 Study design, setting, and participants

This single-center cross-sectional study included consecutive contrast-enhanced VWI examinations performed at Shantou Central Hospital between May 2015 and December 2025. Of 875 examinations, 40 (4.6%) were excluded: 6 because image quality precluded evaluation of the falx free margin, and 34 because of coexisting conditions that could confound dural-enhancement assessment (cranial trauma, intracranial hemorrhage, post-operative changes, or pathological meningeal/meningeal-vascular thickening), leaving 835 evaluable examinations from 811 patients. For every primary analysis, only the first outcome-evaluable examination of each patient was included; the other 24 repeat examinations were reserved for all-examination sensitivity analyses. Exam-level exclusion was non-differential with respect to age (61.7±13.8 vs 60.7±11.8 years; Student t-test, p=0.608), male sex (65.0% vs 62.4%; continuity-corrected chi-square, p=0.869), and emergency presentation (17.5% vs 16.6%; continuity-corrected chi-square, p=1.000). Twenty-two patients contributed two or more evaluable examinations and were included in the repeat-examination reliability analysis.

### 2.2 PISE definition and image reading

PISE was defined operationally as dural enhancement located strictly at the free inferior margin of the falx cerebri, in the peri-ISS dura, whose extent exceeds the contour of the ISS vascular lumen on post-contrast T1-weighted black-blood VWI. All examinations were acquired with three-dimensional T1-weighted black-blood vessel-wall sequences before and after contrast administration. The black-blood preparation of VWI suppresses intraluminal vascular signal, which facilitated the separation of ISS luminal opacification from peri-ISS dural enhancement; comparable black-blood vessel-wall sequences were used in the ISS visualization study of Tsutsumi et al. [7] and in the parasagittal dural study of Joo et al. [13]. Because the ISS lumen itself opacifies in approximately 97% of contrast MRI examinations [7], the reading protocol explicitly separated two fields—ISS luminal opacification and peri-ISS dural enhancement—and PISE was scored only when enhancement extended beyond the luminal contour (tubular or band-like enhancement within the peri-ISS dural space). The thin, linear, discontinuous physiological enhancement described along the falx body, tentorium, and convexity [3–5] lies outside the falx free margin and does not overlap the PISE territory; location restriction was therefore sufficient to distinguish the two. Pericallosal artery opacification was excluded by correlation with angiographic sequences; a persistent falcine sinus (located posteriorly, connecting to the superior sagittal sinus) [8] and falcine calcification were excluded morphologically. PISE is a pachymeningeal (dural) finding; it was distinguished from leptomeningeal (pial) enhancement, which follows cortical sulci and has a different anatomical basis [2]. Examinations showing diffuse supra- and infratentorial smooth dural thickening prompted clinical review for intracranial hypotension [30,31]; nodular or irregular dural thickening prompted consideration of hypertrophic pachymeningitis; and recent surgery or lumbar puncture was recorded because post-procedural meningeal enhancement is common [1,32]. PISE is not part of routine clinical reporting; all examinations were re-read for this study under a pre-specified two-stage protocol. First, before study reading began, a reliability subset of 60 examinations (30 PISE-positive and 30 PISE-negative on preliminary assessment) was randomly selected in advance; within this subset, two readers, each with more than 5 years of professional experience, independently and blindly scored PISE without communication, producing separate datasets. Inter-reader agreement between the two independent blinded readings was almost perfect (Cohen’s κ=0.933, 95%CI 0.842–1.000; observed agreement 96.7%, 58/60), and intra-reader agreement on repeat assessment at a different time was perfect (κ=1.000; 60/60); the two discordant pairs (one positive-to-negative and one negative-to-positive) were resolved by consensus. Second, all 875 examinations—including the reliability subset—were then read jointly by both readers, with discrepancies resolved by consensus, and the consensus values constituted the analytical dataset of this study.

### 2.3 Covariates

Forty-seven variables were abstracted from the same examination episode: demographics (age, sex); vascular risk factors (hypertension, diabetes, hyperlipidemia, hyperuricemia, smoking, alcohol use); care-pathway variables; intracranial atherosclerotic measures on angiographic/vessel-wall imaging—plaque grade (0–2), vulnerable plaque (binary), regional plaques (left anterior, right anterior, posterior circulation), and stenosis degree (0–4); parenchymal and perfusion measures—recent infarction, encephalomalacia, white matter hyperintensity grade (0–3), microbleeds, perfusion abnormality, prolonged time-to-peak (TTP) and mean transit time (MTT); and moyamoya disease.

### 2.4 Statistical analysis

All primary analyses used one first outcome-evaluable examination per patient (n=811). Group comparisons used the chi-square or Fisher exact test for categorical variables and the Mann–Whitney U or Student t-test for continuous/ordinal variables; effect sizes are reported as odds ratios (OR) with 95% confidence intervals or rank-biserial correlations. Two-sided p-values were adjusted across the 47-variable univariate screen using the Benjamini–Hochberg procedure (FDR q<0.05). Candidate variables for multivariable modelling were restricted to FDR-significant variables with less than 15% missingness; age and sex were forced into the model; the perfusion-imaging series (≈17% missing) was thereby excluded from multivariable modelling but retained in univariate reporting. Backward elimination was applied (retention p<0.05 and variance inflation factor [VIF] <10; no variable required VIF removal). The complete-case model comprised n=732 with 563 events (events-per-variable 31.3, based on 18 candidate variables). Age was modelled per +1 SD, and plaque grade and stenosis degree per +1 grade. Internal validation used 5-fold patient-grouped cross-validation, with patient identifier as the grouping variable and no patient shared between training and test folds. Univariate screening, FDR correction, missingness filtering, backward elimination, and age standardization were repeated within each training fold; the validation estimand was the complete modelling pipeline. Pooled out-of-fold (OOF) AUCs were reported with 95% CIs from 1,000 patient-cluster bootstrap resamples. LASSO logistic regression used patient-grouped internal cross-validation with fold-specific scaling [33]; random forest used patient-grouped 5-fold cross-validation, out-of-fold permutation importance, and partial dependence analysis [34]. Dose–response across plaque grades was tested with the Cochran–Armitage trend test. Circulation-specific associations were estimated in two logistic models adjusted for age and sex: a regional model entering right anterior, left anterior, and posterior-circulation plaque simultaneously, and a combined model entering any anterior-circulation plaque versus posterior-circulation plaque. Sensitivity analyses comprised (S1) simple imputation with the full patient sample (n=811), (S2) multiple imputation (5 datasets, Rubin pooling), (S3) repeating the final-variable model using all evaluable examinations (n=791 complete cases), and univariate re-testing using all 835 evaluable examinations. An exploratory quantification of association strength for vulnerable plaque compared receiver-operating-characteristic areas under the curve (AUC) for age+sex versus age+sex+PISE. Analyses used R software (version 4.2.3) and Python (version 3.12.3). Reporting follows the STROBE guideline for cross-sectional studies [35].

## 3. Results

### 3.1 Cohort and PISE detection rate

Of 875 consecutive contrast-enhanced VWI examinations, 835 were evaluable (40 excluded: 6 for image quality and 34 for coexisting conditions confounding dural-enhancement assessment; non-differential exclusion as detailed in Methods), corresponding to 811 patients. In the patient-level primary analysis, PISE was detected in 609/811 patients (75.1%); 202/811 (24.9%) were PISE-negative. Detection rose monotonically with age: 50.0% (22/44) in those <40 years, 70.0% (196/280) at 40–60 years, 79.3% (230/290) at 60–70 years, and 81.7% (161/197) in those >70 years (logit-linear trend p=5.6×10⁻⁶; OR=1.55 per ascending age stratum) (Figure 2). Twenty-two patients had two or more evaluable examinations (median interval 206 days between the first and last examination). PISE status was concordant in 16 (73%); all six discordant pairs showed incident PISE at follow-up (0→1), accompanying stable or increased atherosclerotic burden or older age—an exploratory observation consistent with temporality between burden accumulation and sign emergence. In the pre-specified reliability subset (60 examinations, two independent blinded readers), inter-reader agreement for PISE was almost perfect (κ=0.933, 95%CI 0.842–1.024; 96.7%, 58/60), and intra-reader agreement was perfect (κ=1.000, 60/60); consensus values from dual reading were used for all analyses.

**Figure 2.**
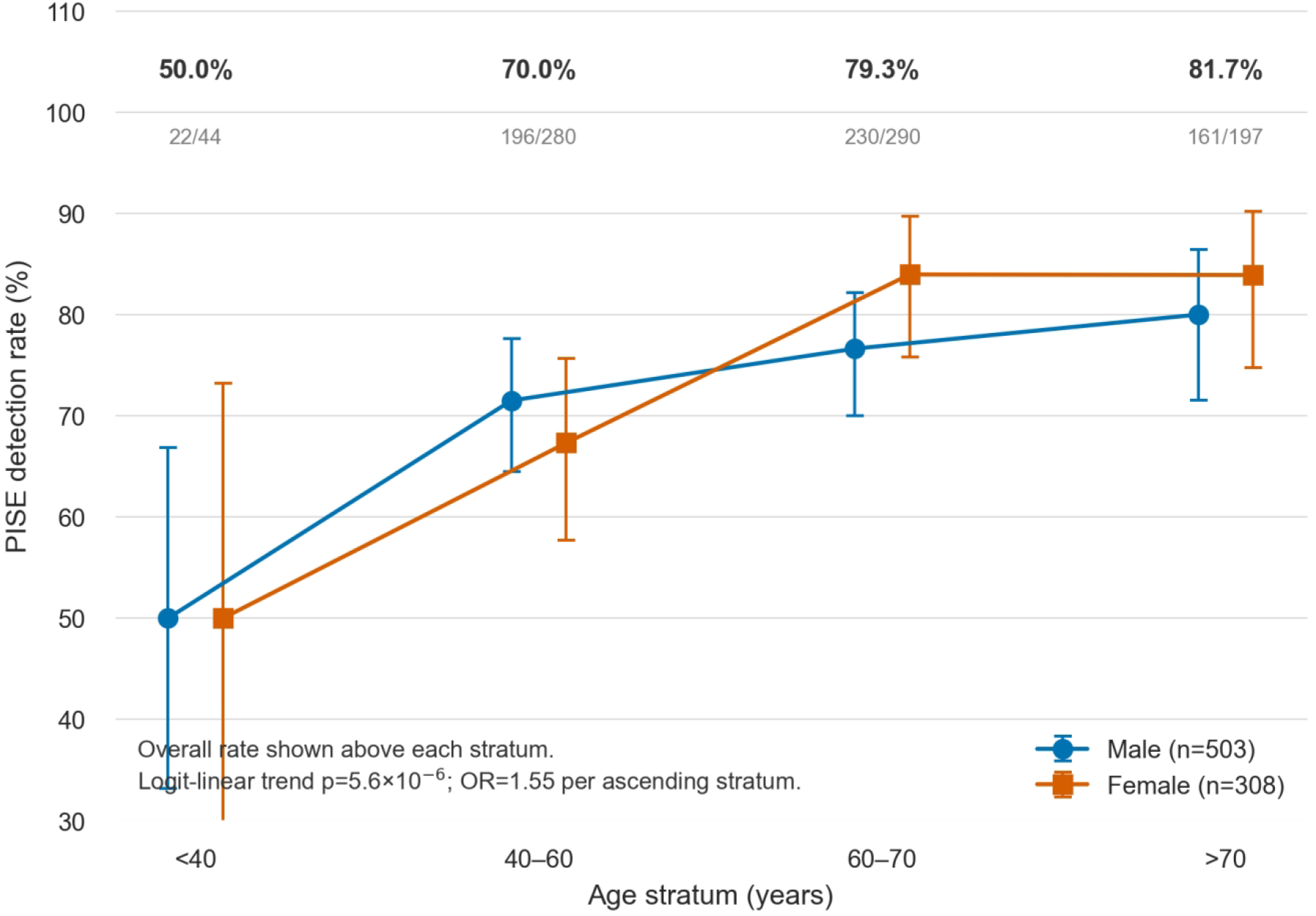
PISE detection rate rises monotonically with age in both sexes. Points and connecting lines show the PISE detection rate by age stratum for male (n=503) and female (n=308) patients; error bars indicate 95% Wilson score confidence intervals; the overall detection rate and n/N are printed above each stratum. Detection rose from 50.0% (22/44) in patients <40 years to 81.7% (161/197) in those >70 years (logit-linear trend p=5.6×10⁻⁶; OR=1.55 per ascending age stratum; 811 evaluable patients). Abbreviations: CI, confidence interval; OR, odds ratio; PISE, para-ISS dural space enhancement.

### 3.2 Baseline characteristics

Median age was 63 years (IQR 54–69); 62.0% were male and 93.7% of patients with recorded encounter type were inpatient (Table 1). Compared with PISE-negative patients, PISE-positive patients were older (median 64 [56–70] vs 59 [48.25–67] years; p=1.1×10⁻⁶, q=8.6×10⁻⁶), more often inpatient (95.9% vs 87.2%; q=2.8×10⁻⁴), and more often emergency referrals (19.5% vs 9.9%; q=0.008). Among vascular risk factors, hypertension history survived FDR correction (84.0% vs 75.7%; q=0.043), whereas diabetes (40.9% vs 33.0%; q=0.126), hyperlipidemia (70.6% vs 65.9%; q=0.363), and hyperuricemia (25.9% vs 18.5%; q=0.126) did not; smoking history was less frequent among PISE-positive patients but did not survive FDR correction (21.3% vs 29.3%; q=0.091). Sex distribution did not differ (p=0.383).

**Table 1.** PISE-positive patients are older and more often inpatient and emergency referrals. Data are median [IQR] or n/N (%). P-values are from the Mann–Whitney U test (age) or continuity-corrected chi-square tests (categorical variables); q = Benjamini–Hochberg FDR-adjusted p-value across the 47-variable screen. Abbreviations: FDR, false discovery rate; IQR, interquartile range; PISE, para-ISS dural space enhancement.

| **Variable** | **Overall (n=811)** | **PISE-positive (n=609)** | **PISE-negative (n=202)** | **Test** | **p** | **q (FDR)** |
|---|---|---|---|---|---|---|
| Age (years), median [IQR] | 63[54,69] | 64.00[56.00,70.00] | 59.00[48.25,67.00] | Mann-Whitney U | 1.1×10<sup>−6</sup> | 8.6×10<sup>−6</sup> |
| Male sex, n/N (%) | 503/811 (62.0%) | 372/609 (61.1%) | 131/202 (64.9%) | Chi-square | 0.383 | 0.439 |
| Inpatient encounter, n/N (%) | 719/767 (93.7%) | 556/580 (95.9%) | 163/187 (87.2%) | Chi-square | 4.2×10<sup>−5</sup> | 2.8×10<sup>−4</sup> |
| Emergency presentation, n/N (%) | 139/811 (17.1%) | 119/609 (19.5%) | 20/202 (9.9%) | Chi-square | 0.002 | 0.008 |
| Smoking history, n/N (%) | 164/711 (23.1%) | 118/554 (21.3%) | 46/157 (29.3%) | Chi-square | 0.046 | 0.091 |
| Alcohol use history, n/N (%) | 61/709 (8.6%) | 45/553 (8.1%) | 16/156 (10.3%) | Chi-square | 0.502 | 0.548 |
| Hypertension history, n/N (%) | 601/732 (82.1%) | 473/563 (84.0%) | 128/169 (75.7%) | Chi-square | 0.019 | 0.043 |
| Diabetes history, n/N (%) | 289/741 (39.0%) | 231/565 (40.9%) | 58/176 (33.0%) | Chi-square | 0.073 | 0.126 |
| Hyperlipidemia, n/N (%) | 481/692 (69.5%) | 373/528 (70.6%) | 108/164 (65.9%) | Chi-square | 0.286 | 0.363 |
| Hyperuricemia, n/N (%) | 166/688 (24.1%) | 136/526 (25.9%) | 30/162 (18.5%) | Chi-square | 0.071 | 0.126 |

*Table note. Data are median [IQR] or n/N (%). P-values are from the Mann–Whitney U test (age) or continuity-corrected chi-square tests (categorical variables); q = Benjamini–Hochberg FDR-adjusted p-value across the 47-variable screen. Abbreviations: FDR, false discovery rate; IQR, interquartile range; PISE, para-ISS dural space enhancement.*

### 3.3 Univariate association spectrum

Of 47 variables screened, 21 were significant after FDR correction (q<0.05) (Figure 3). The strongest signals came from the atherosclerotic burden spectrum: vulnerable plaque (90.9% vs 54.5%; OR=8.29, 95%CI 5.54–12.40; q<1×10⁻¹⁵), plaque grade (r=0.478; q<1×10⁻¹⁵), left anterior plaque (OR=2.85, 2.00–4.07), right anterior plaque (OR=2.67, 1.87–3.83), posterior-circulation plaque (OR=2.41, 1.70–3.42), and stenosis degree (r=0.158; q=0.002). A second cluster comprised parenchymal and perfusion alterations: encephalomalacia (OR=3.11, 1.40–6.92), perfusion abnormality (OR=2.11, 1.34–3.34), prolonged TTP (OR=2.02, 1.31–3.11), microbleeds (OR=1.96, 1.33–2.90), prolonged MTT (OR=1.82, 1.18–2.80), recent infarction (OR=1.92, 1.39–2.65), posterior-circulation infarction (OR=1.83, 1.14–2.93), absence of perfusion deficit on perfusion imaging (OR=1.69, 1.12–2.55), lacunar lesions (OR=1.64, 1.10–2.44), and higher white matter hyperintensity grade (r=0.172; q=3.1×10⁻⁴). A third cluster comprised demographic, clinical-history, and care-pathway variables: inpatient status (OR=3.41, 1.89–6.17), emergency presentation (OR=2.21, 1.34–3.66), hypertension history (OR=1.68, 1.11–2.56; q=0.043), and age (r=0.228; q=8.6×10⁻⁶). Moyamoya disease was inversely associated (1.2% vs 4.5%; OR=0.25, 0.09–0.68; Fisher exact; q=0.020), serving as a natural control.

**Figure 3.**
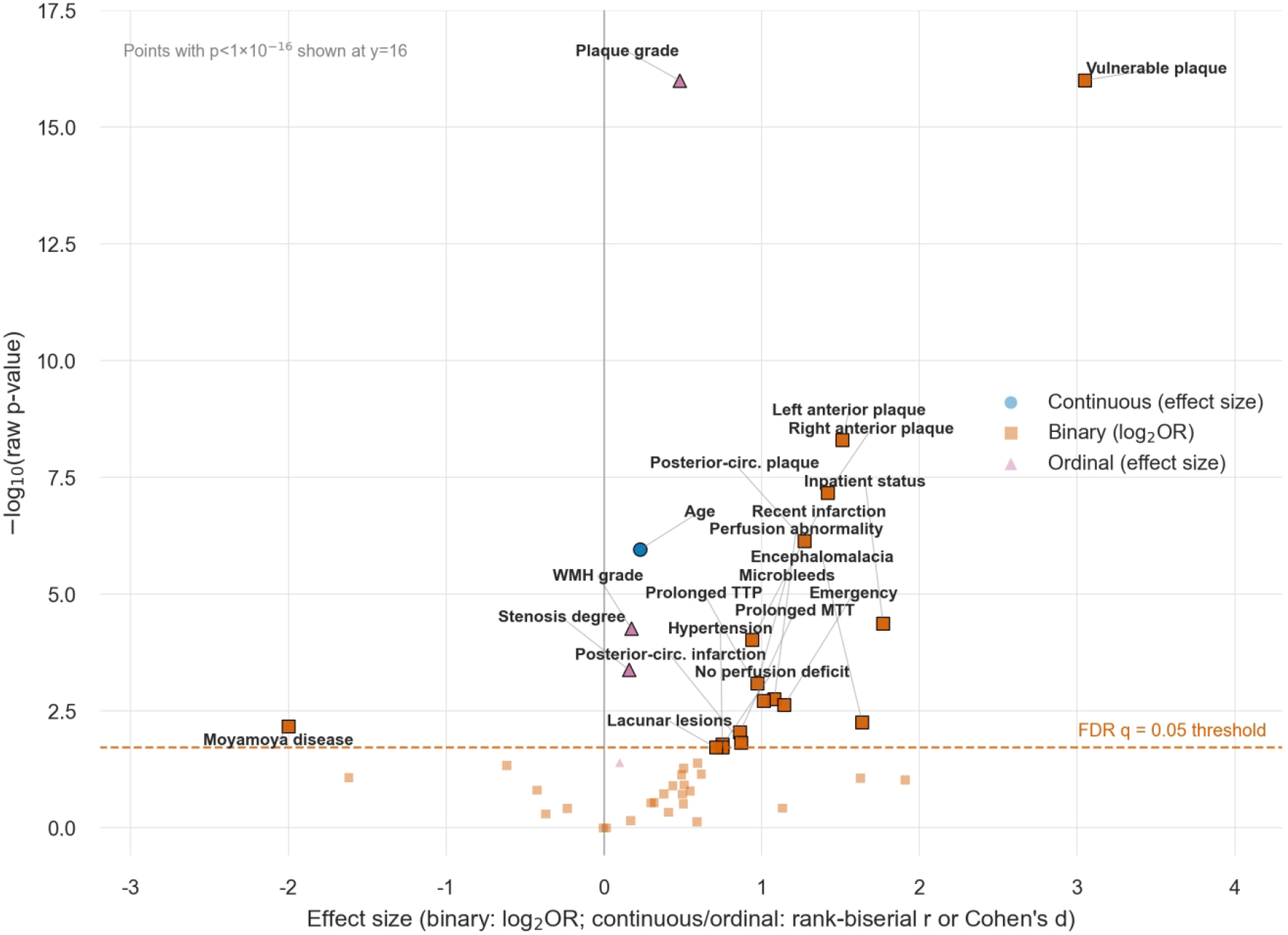
Atherosclerotic burden variables dominate the univariate association spectrum of PISE. Volcano plot of the 47 demographic, clinical-history, and imaging variables screened univariately in the patient-level primary analysis; x-axis, effect size (binary variables: log₂ odds ratio; continuous/ordinal variables: rank-biserial correlation or Cohen’s d); y-axis, −log₁₀(raw p-value). The dashed line marks the Benjamini–Hochberg FDR q=0.05 threshold; the 21 FDR-significant variables are outlined in black and labeled; points with p<1×10⁻¹⁶ are displayed at y=16. Moyamoya disease is the sole inverse FDR-significant association. Abbreviations: FDR, false discovery rate; MTT, mean transit time; OR, odds ratio; PISE, para-ISS dural space enhancement; TTP, time-to-peak; WMH, white matter hyperintensity.

### 3.4 Dose–response and anatomical correspondence

PISE detection increased stepwise with plaque grade: 38.7% (53/137) for grade 0, 77.2% (247/320) for grade 1, and 90.3% (280/310) for grade 2 (Cochran–Armitage Z=11.09, p<1×10⁻¹⁵; n=767 evaluable patients) (Figure 4). In the regional model adjusted for age and sex, right anterior plaque (OR=2.74, 1.86–4.05; p=4.23×10⁻⁷), left anterior plaque (OR=2.95, 2.00–4.34; p=4.20×10⁻⁸), and posterior-circulation plaque (OR=2.49, 1.69–3.68; p=4.48×10⁻⁶) were each associated with PISE. In the combined model, anterior-circulation plaque showed a stronger association (OR=4.42, 3.06–6.39; p=2.84×10⁻¹⁵) than posterior-circulation plaque (OR=2.76, 1.86–4.10; p=5.25×10⁻⁷)—an approximately 1.6-fold ratio consistent with the anterior-circulation deep drainage territory of the ISS.

**Figure 4.**
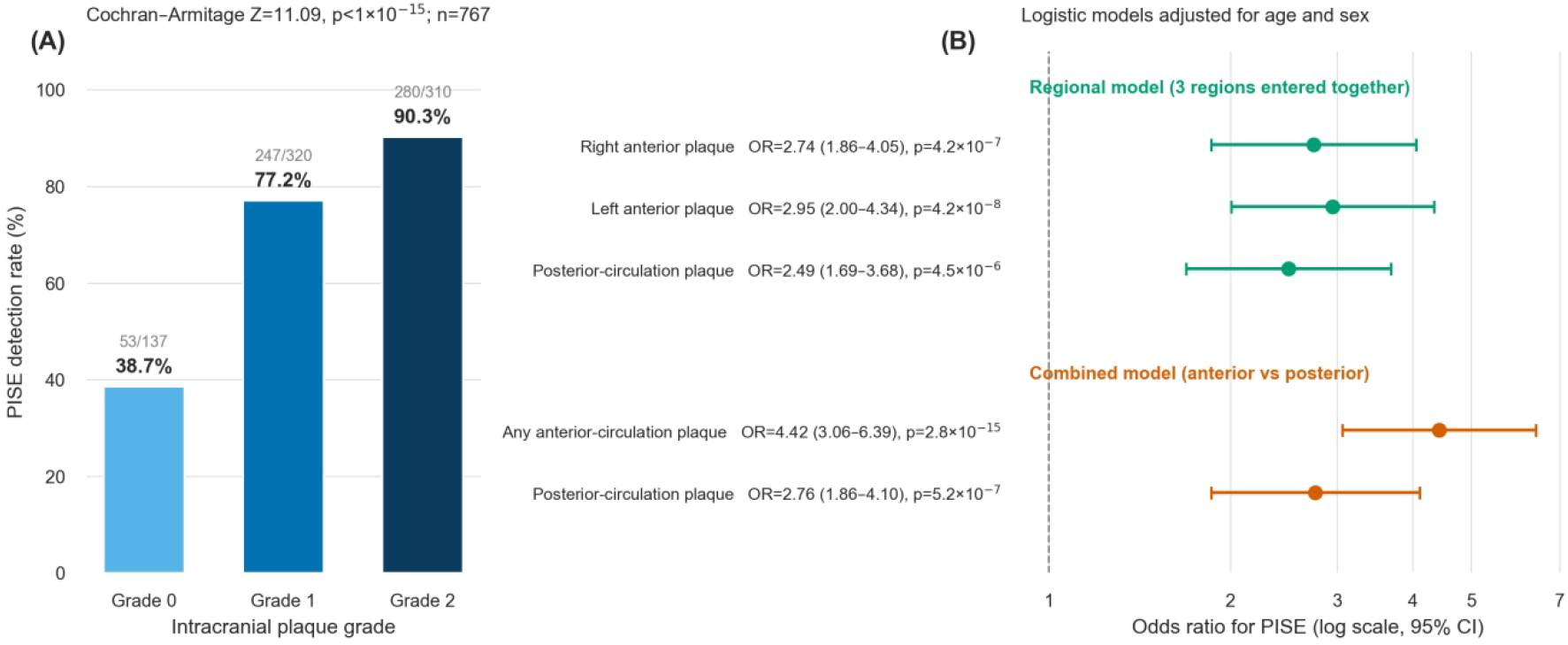
PISE detection shows a monotone dose–response across plaque grades and an anterior-predominant anatomical correspondence. (A) PISE detection rate across intracranial plaque grades 0, 1, and 2 (38.7% [53/137], 77.2% [247/320], and 90.3% [280/310]); error bars indicate 95% Wilson score confidence intervals; the trend was tested with the Cochran–Armitage test (Z=11.09, p<1×10⁻¹⁵; n=767 patients with evaluable plaque grade). (B) Forest plot of circulation-specific associations from logistic models adjusted for age and sex: a regional model entering right anterior, left anterior, and posterior-circulation plaque simultaneously, and a combined model entering any anterior-circulation versus posterior-circulation plaque (OR=4.42 vs 2.76); points show odds ratios and horizontal lines show 95% confidence intervals on a log scale; OR (95% CI) and exact p-values are listed at right. Abbreviations: CI, confidence interval; OR, odds ratio; PISE, para-ISS dural space enhancement.

### 3.5 Multivariable model

The final multivariable model retained six variables (Figure 5): vulnerable plaque (OR=2.37, 1.07–5.22; p=0.033), plaque grade (OR=2.84 per grade, 1.77–4.57; p=1.6×10⁻⁵), age (OR=1.38 per SD≈11.4 years, 1.15–1.66; p=7.1×10⁻⁴), and inpatient status (OR=3.18, 1.05–9.61; p=0.041) as positive independent correlates; stenosis degree (OR=0.83 per grade, 0.69–0.99; p=0.035) as an inverse correlate; and male sex (forced in; OR=0.76, 0.51–1.15; p=0.194, not significant). Apparent AUC was 0.775, with good calibration (Hosmer–Lemeshow p=0.670); patient-grouped 5-fold pooled OOF AUC for the complete modelling pipeline was 0.755 (95%CI 0.713–0.799, patient-cluster bootstrap; valid OOF n=767). LASSO with patient-grouped cross-validated penalty selection retained five variables (plaque grade, age, vulnerable plaque, inpatient status, and microbleeds), including all four positive independent correlates of the main model, supporting model stability. The random forest did not outperform the logistic modelling pipeline (patient-grouped OOF AUC=0.742, 95%CI 0.696–0.787; valid OOF n=760), suggesting predominantly linear/additive associations; its out-of-fold permutation importance ranking (plaque grade 0.0433, age 0.0287, vulnerable plaque 0.0186) mirrored the logistic model, and partial dependence plots showed a high plateau of PISE probability beyond approximately 55 years and a near-linear increase across plaque grades. As an exploratory quantification of association strength, PISE alone discriminated vulnerable plaque with AUC=0.727 (sensitivity 0.838, specificity 0.616, positive predictive value 0.909, negative predictive value 0.455, accuracy 0.798; vulnerable plaque frequency 0.820), and adding PISE to age+sex raised the AUC from 0.619 to 0.770 (ΔAUC=+0.151; PISE coefficient 2.056, p=2.80×10⁻²²).

**Figure 5.**
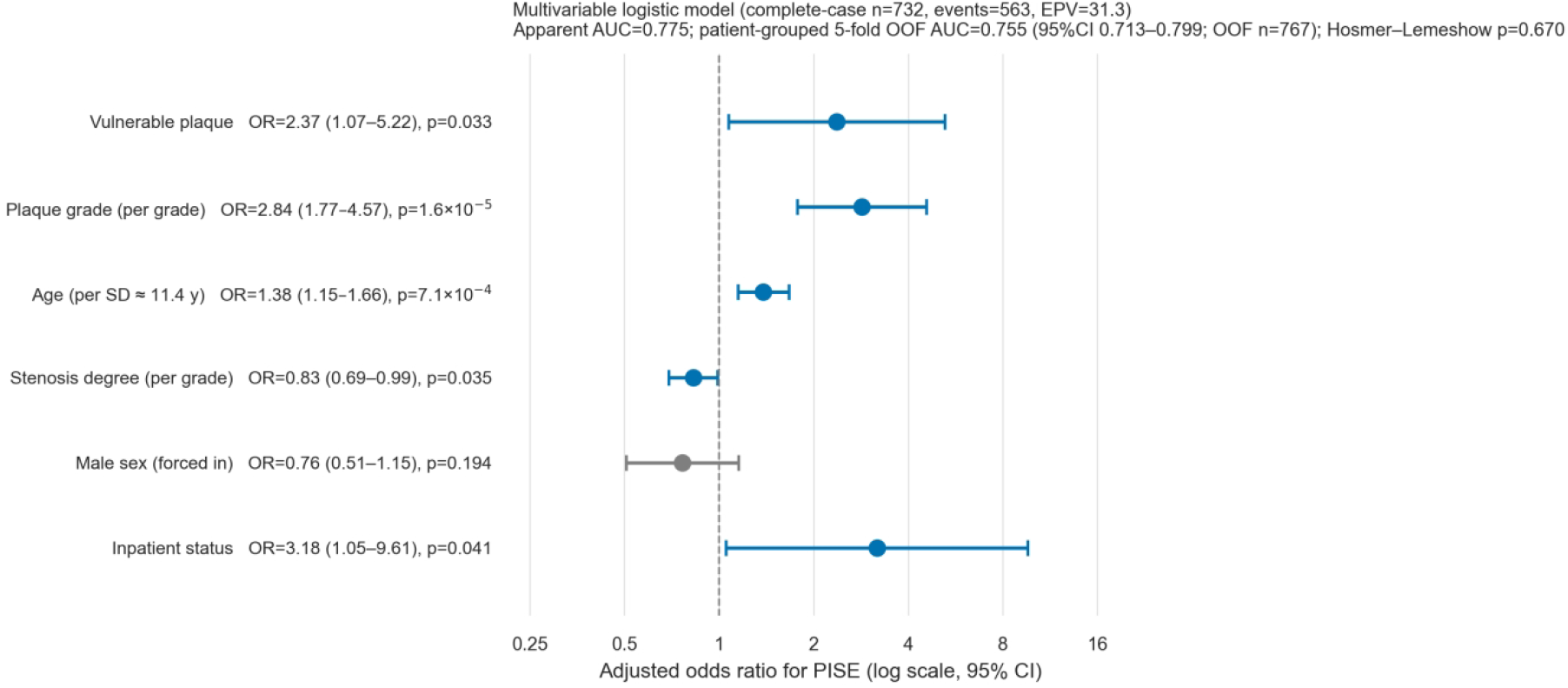
Vulnerable plaque, plaque grade, age, and inpatient status are independent positive correlates of PISE. Forest plot of the six variables retained in the final multivariable logistic regression model (complete-case n=732, events=563, events-per-variable=31.3); points show adjusted odds ratios and horizontal lines show 95% confidence intervals on a log scale; variables significant at p<0.05 are shown in blue and the non-significant forced-in covariate (male sex) in gray. OR for age is per +1 SD (1 SD≈11.4 years); ORs for plaque grade and stenosis degree are per +1 grade. Model performance: apparent AUC=0.775; patient-grouped 5-fold pooled OOF AUC=0.755 (95% CI 0.713–0.799; valid OOF n=767); Hosmer–Lemeshow p=0.670. Abbreviations: AUC, area under the curve; CI, confidence interval; OR, odds ratio; PISE, para-ISS dural space enhancement; SD, standard deviation.

### 3.6 Sensitivity analyses

All sensitivity analyses were directionally concordant with the main model. With simple imputation in the full patient sample (n=811), age (OR=1.37, p=2.9×10⁻⁴) and plaque grade (OR=3.03, p=1.7×10⁻⁶) remained significant; vulnerable plaque (OR=1.93, p=0.077), inpatient status (OR=1.66, p=0.155), and stenosis degree (OR=0.86, p=0.056) were not significant. With multiple imputation, age (OR=1.37, p=4.0×10⁻⁴), plaque grade (OR=2.44, p=5.7×10⁻⁴), vulnerable plaque (OR=2.69, p=0.014), and stenosis degree (OR=0.83, p=0.030) remained significant, whereas inpatient status did not (OR=1.70, p=0.147). In the all-examination final-variable model (n=791 complete cases, 602 events; apparent AUC=0.780), age (OR=1.45), plaque grade (OR=2.74), vulnerable plaque (OR=2.53), and stenosis degree (OR=0.82) remained significant (all p<0.05), whereas inpatient status did not (OR=1.55, p=0.203). When all 835 evaluable examinations were used for univariate sensitivity analysis, all 21 patient-level FDR-significant findings remained FDR-significant after Benjamini–Hochberg re-correction across the 47-variable set (maximum q=0.048).

## 4. Discussion

In 811 patients with evaluable contrast-enhanced VWI, we defined and characterized a previously undescribed dural enhancement sign—para-ISS dural space enhancement (PISE)—detected in 75.1% of this indication-enriched cohort. Four mutually reinforcing lines of evidence link PISE to intracranial atherosclerotic burden: (i) independent association after multivariable adjustment (vulnerable plaque OR=2.37; plaque grade OR=2.84 per grade; age OR=1.38 per SD; inpatient status OR=3.18); (ii) a monotone dose–response across plaque grades (38.7%→77.2%→90.3%; Z=11.09); (iii) anatomical correspondence, with anterior-circulation plaque more strongly associated than posterior-circulation plaque (OR=4.42 vs 2.76); and (iv) a natural control—moyamoya disease, a non-atherosclerotic arteriopathy, showed markedly lower detection (OR=0.25). The findings were consistent across imputation and all-examination sensitivity analyses. The longitudinal observation that all six discordant repeat-examination pairs showed incident PISE at follow-up—never regression—adds a temporal dimension consistent with burden accumulation preceding sign emergence, although it derives from only 22 patients.

Why should enhancement concentrate at this particular site? The concentration of the sign at the falx free margin has a coherent anatomical logic. The ISS drains the falx itself and the medial hemispheric deep territory of the anterior circulation toward the vein of Galen and straight sinus [6], and histological work has confirmed lymphatic vessels accompanying the ISS in human dura [16]. The arterial supply of the anterior and inferior falx derives from anterior-circulation sources—the anterior falcine artery (a meningeal branch of the anterior ethmoidal artery [6]) and distal dural branches of the anterior cerebral artery—demonstrated angiographically and anatomically [36,37]. PISE therefore sits at a functional junction where anterior-circulation deep drainage meets the dura. This anatomy predicts precisely what we observed: anterior-circulation plaque associated more strongly than posterior-circulation plaque. We do not claim that the ISS is a unique lymphatic site—dural lymphatics are widely distributed [16,18]—but that the peri-ISS dura is a plausible hub for anterior-circulation drainage–dural interface signaling, a hub that published 7T atlases happened not to sample [19].

The correlate pattern constrains mechanism in an informative way. PISE was associated with established vascular lesion burden—plaque grade, plaque vulnerability, and downstream parenchymal/perfusion injury—whereas most risk-factor exposures (diabetes, hyperlipidemia, hyperuricemia) did not survive multiple-testing correction and hypertension history was only modestly associated (OR=1.68; q=0.043). The sign thus tracks end-organ disease rather than risk-factor exposure, and it does so in a graded fashion. One parsimonious interpretation is a dural drainage-interface response: atherosclerotic and parenchymal brain injury increases interstitial solute and fluid load, which is cleared partly through glymphatic and intramural peri-arterial routes toward meningeal lymphatic and parasagittal dural compartments [9,10,12,20,22]; sustained load at the peri-ISS interface could remodel the local dura (vascularity, permeability, or lymphatic caliber), rendering it visibly enhancing on contrast-enhanced VWI. This hypothesis is compatible with evidence that CSF tracer effluxes to parasagittal dura [12], that parasagittal dural kinetics change with age [13–15], that experimental augmentation of dural lymphatics alters ischemic brain outcomes [38], and that the parasagittal dura functions as a neuroimmune interface [21]. The high background detection rate should be read together with the physiological-enhancement literature: PISE may represent a pathological amplification of a physiological phenomenon (a graded, load-related continuum) rather than an all-or-none abnormality; the dose–response gradient argues against a fixed anatomical variant. We explicitly refrain from claiming exclusion of alternative mechanisms, which requires targeted prospective designs.

The inverse association in moyamoya disease (OR=0.25) is mechanistically informative. Moyamoya displays prominent leptomeningeal change—the ivy sign of slow pial collateral flow on FLAIR and post-contrast images [39]—yet spontaneous pachymeningeal enhancement is not a feature; indeed, all six moyamoya patients in the delayed vessel-wall MRI series of Umehana et al. showed no dural enhancement outside postoperative areas [24]. This leptomeningeal–pachymeningeal dissociation indicates that PISE is neither a reflection of pial slow flow nor of collateralization per se. Within our framework, terminal anterior-circulation occlusion in moyamoya reshapes deep drainage demand and pathways, plausibly reducing load at the peri-ISS dural interface—the mirror image of the atherosclerotic-burden association.

The closest published observation is the delayed ipsilateral dural/meningeal enhancement after large cerebral infarction reported by Umehana et al. [24]. The two phenomena are best regarded as complementary rather than identical, differing on four dimensions: theirs is a delayed-phase (1–6 h post-injection) vessel-wall MRI finding, triggered by acute large infarction, distributed over the ipsilateral cortical dura, and described in 11 patients, whereas PISE is detected at standard post-contrast timing on VWI, tracks chronic atherosclerotic burden, is centered on the midline ISS at the falx free margin, and was assessed in 811 patients. Their compensatory-drainage interpretation after acute drainage-route injury is conceptually aligned with the chronic load-related interface response we propose.

To our knowledge, this is the first study to define an ISS-centered dural enhancement sign and to examine its correlates systematically. More importantly, the coherence of the clinical and anatomical findings, together with their robustness across complementary analytical approaches, supports the interpretation of PISE as a reproducible imaging marker associated with vascular burden rather than an incidental enhancement pattern. Because PISE can be assessed on the same VWI examination used for plaque evaluation, it could be incorporated into existing imaging workflows without additional scanning.

Several limitations qualify these findings. First, the study was primarily cross-sectional; exploratory repeat examinations in 22 patients provided preliminary temporal context for the emergence of PISE, but were insufficient to support robust temporal or causal inference. Second, the cohort is a single-center, indication-enriched clinical spectrum (93.7% inpatients); the 75.1% figure is a detection rate within this spectrum and is not generalizable to the general population. Third, VWI is not a lymphatic-specific sequence, so the drainage-interface mechanism remains inferential; delayed-phase and DCE sub-studies are needed; moreover, the visibility of PISE on conventional post-contrast T1-weighted sequences has not been verified, and extrapolation of the present detection rate to non-VWI protocols should be made with caution. Finally, the inverse association of stenosis degree with PISE should be interpreted cautiously given its divergent univariable result.

## 5. Conclusions

The para-ISS dural space enhancement (PISE) sign is a frequently detected, previously undescribed dural enhancement at the falx free margin on contrast-enhanced intracranial vessel-wall MRI. It is independently associated with intracranial atherosclerotic burden, with a monotone dose–response across plaque grades and an anterior-predominant anatomical correspondence that matches ISS drainage territory, while the non-atherosclerotic arteriopathy moyamoya disease shows markedly lower detection. The correlate pattern is compatible with an atherosclerosis-related dural drainage-interface response. Prospective studies with dedicated delayed/DCE sequences, formal reliability testing, and histological correlation are warranted to validate the sign and test the proposed mechanism.

## Declarations

### Ethics approval and consent to participate

This study was approved by the Ethics Committee of Shantou Central Hospital (approval number: [2026] Research No. 049). The requirement for informed consent was waived.

### Data availability

Data are available from the corresponding author upon reasonable request.

